# A catalogue of missense and nonsense mutation abundances for the U.S. cancer patient population

**DOI:** 10.64898/2026.04.20.26351248

**Authors:** Adith S. Arun, David Liarakos, Gaurav Mendiratta, Thomas McFall, Diana C. Hargreaves, Geoffrey M. Wahl, Jiancheng Hu, Edward C. Stites

## Abstract

Widespread genomic sequencing efforts have characterized the molecular foundations of the different cancers. By combining these genomic data in a manner proportional to the population-level abundances of these different cancers, we estimate the overall abundances of each observed missense and nonsense mutation within the U.S. cancer patient population. We find *BRAF* V600E (5.2%) is the most common mutation in the cancer patient population, *TP53* R175H (1.5%) is the most common tumor suppressor mutation, and *APC* R876X (0.4%) is the most common nonsense mutation. These values differ largely and significantly from what would be found in a typical pan-cancer analysis, where different cancer types are included out of proportion to population level incidence. We present the full ordered lists of population-level abundances for specific missense and nonsense mutations, and we demonstrate the value of these data by further analyzing high priority genes (e.g., *TP53*, *KRAS*, *BRAF*) and pathways (e.g., RTK/RAS, PI3K, and WNT/β-catenin). Overall, this information is a resource that should benefit the basic science, translational, and clinical cancer research communities.

## INTRODUCTION

Aggregated cancer genomic data, as are found in published pan-cancer analyses (*1–4*) and cancer genomic databases (*5–7*), have provided informative portrayals of cancer biology across many forms of cancer. The aggregate proportion of samples with a mutation in these pan-cancer databases and resources has commonly been used as a surrogate for the population level proportion of cancer patients with that mutation (*8*). This is problematic, in part, because the proportions of the different cancers in these databases and resources are generally not the same as the proportions of individuals with that form of cancer for any specific population.

One approach for obtaining population-level estimates for the frequency of patients with mutations in a given gene (or for the proportion of patients with a specific mutation) involves reweighting the cancer-type mutation frequencies by the population-level, cancer-type proportions. Such data can be found in public health and epidemiological resources (*9*). Reweighting the contributions of different subsets of data to the desired proportion is an effective approach to correcting for biases within large datasets (*10*). Two challenges have complicated the reweighting of cancer genomic data to better represent a specific population. First, although cancer epidemiological data, like that from the National Cancer Institute (NCI) Surveillance, Epidemiology, and End Results (SEER) cancer epidemiology data repository (*9*), commonly utilize a formalized approach for characterizing cancer, like World Health Organizations (WHO) International Classification of Diseases for Oncology, 3^rd^ edition (ICD-O-3) descriptive codes (*11*), cancer genomics has frequently communicated their studies using ad hoc and colloquial cancer descriptions (*12*). Because of these different approaches to nomenclature, cancer genomic data and epidemiological data cannot be directly integrated in their current forms. Second, different cancer genomic studies that claim to have studied the same type of cancer often have different inclusion criteria. For example, some pancreas cancer exome data sets might only include pancreatic adenocarcinoma (*13, 14*) whereas others would include pancreatic adenocarcinoma as well as other tumors of the pancreas, such as pancreatic neuroendocrine cancer (*4, 15*). Similarly, some studies of melanoma might only include cutaneous melanoma (*16, 17*), while another includes cutaneous and acral melanoma (*4*) and another includes cutaneous, acral, and uveal melanoma (*18*). Thus, although there are efforts to formalize cancer nomenclature as it has been utilized within cancer genomics through the OncoTree (*12*), simple mapping to the OncoTree will not suffice due to these different inclusion criteria.

To overcome these challenges in cancer genomics, we previously developed an integrative nomenclature re-mapping approach we termed ROSETTA (Reclassification Of Sequencing and Epidemiological Tumor Type Annotations) (*19, 20*). ROSETTA enables translation of nomenclatures between terms used in epidemiology and genomics to a common set of cancer ROSETTA classifications. Additionally, our analysis workflow includes mapping individual cancer samples within different genomics studies to specific ROSETTA classifications based on the metadata associated with those samples, thus overcoming the variations in inclusion criteria.

In our foundational work developing ROSETTA, we applied the workflow to analyze gene-level mutation burdens within the population of patients with a new malignant cancer diagnosis within the United States. The data from that study clarified misperceptions about the prevalence of the most abundant oncogenes and has been a useful resource. These data also have public health implications, such as by demonstrating the degree to which cancer research efforts on specific cancer driver genes are aligned to the relative frequency of those genes being mutated in cancer (*21*). The ROSETTA pipeline can also be adapted to new purposes, such as the curation of population level acquired cysteine mutations to evaluate the abundances of these potential drug targets (*22*).

Our original integration of genomics and epidemiology to generate improved estimates of mutation abundances within incident cancers focused on the gene level (*19*). For example, that study projected 35% of new malignant cancer diagnoses include a *TP53* mutation and projected that only 14% of new malignant cancer diagnoses harbor at least one mutation in one of the three *RAS* genes (*KRAS*, *NRAS*, and *HRAS*) which was less than half of the 33% *RAS* mutation rate commonly stated in the literature (*23, 24*). Although gene-level mutation abundance data is useful, it is well appreciated that different mutations to the same gene may have different functional consequences and may respond differently to the same treatment (*25–27*). Additionally, specific missense mutations may serve as drug targets and/or as indications for FDA approved drugs (*28, 29*). Many variants to a known driver gene may be as yet uncharacterized and thus be better considered a variant of unknown significance; some may even be passenger mutations with no functional consequence. Thus, there are many benefits to an analysis of specific mutations.

In the present study, we adapted our workflow for the goal of characterizing the population level burden of individual missense and nonsense mutations. We demonstrate that the implementation of epidemiologically-informed corrections results in proportionately large and significant changes from the pooled exome data. We focus our analysis on recurrent missense and nonsense mutations in established cancer driver genes. We characterize the most common mutations overall, the most common mutations for high importance genes, the most common mutations for high importance pathways, and the distribution between different classes of mutations for high importance genes. In addition to providing the hierarchy of individual mutations in terms of overall numbers of cancer patients effected, this analysis also highlights large variations in contextual influence – with some drivers only being observed in one or a few cancers and others occurring broadly. Overall, these data should provide a more accurate portrayal of cancer mutation abundances than common, cancer-type biased, pan-cancer analyses. These data additionally characterize the average approximate magnitude of error at 35% within the raw, non-corrected, pan-cancer cohort. The useful resource that we present should have a wide set of applications in the basic science, translational, and clinical cancer research communities.

## RESULTS

### The generation of epidemiologically corrected gene mutation frequencies

We utilized a cohort of cancer samples from 140 different, publicly available, cancer exome and genome studies. The cohort comprises 24,431 different cancer exomes and genomes from the same number of unique patients. In our previous work, we manually curated these samples into different ROSETTA cancer-type classification codes on the basis of available metadata to enable integration with epidemiological data (*19*). To aggregate the genomic data in a manner proportional to U.S. cancer incidence rates, we utilized data from the NCI-SEER (*9*) that we converted from WHO ICD-O-3 cancer-type codes (*11*) to ROSETTA cancer-type codes. The re-mapped epidemiological incidence data for each cancer type were then used to reweight the corresponding cancer-type specific rates of mutations for each specific missense or nonsense mutation within the cohort.

Across all exome studies, 2.11 million missense and nonsense mutations were called. This total is from 1.96 million distinct missense mutations in 21,316 different genes and from 144,509 distinct nonsense mutations in 18,390 different genes. Of these missense and nonsense mutations, 203,047 were recurrent mutations observed in two or more patients.

The proportions of different forms of cancer within our annotated cohort are clearly not the same as the cancer type population-level proportions (**Figure 1A**). As opposed to traditional pan-cancer analyses, we removed this bias in representation of the forms of cancer by reweighting each sample from each form of cancer so that the contributions to the entire collection are proportional to the epidemiological incidence (**Supplementary Figure 1**). This results in strict agreement between the proportional representation of each cancer type within the reweighted genomic data and the epidemiological data (**Figure 1A, inset**).

**Figure 1.**
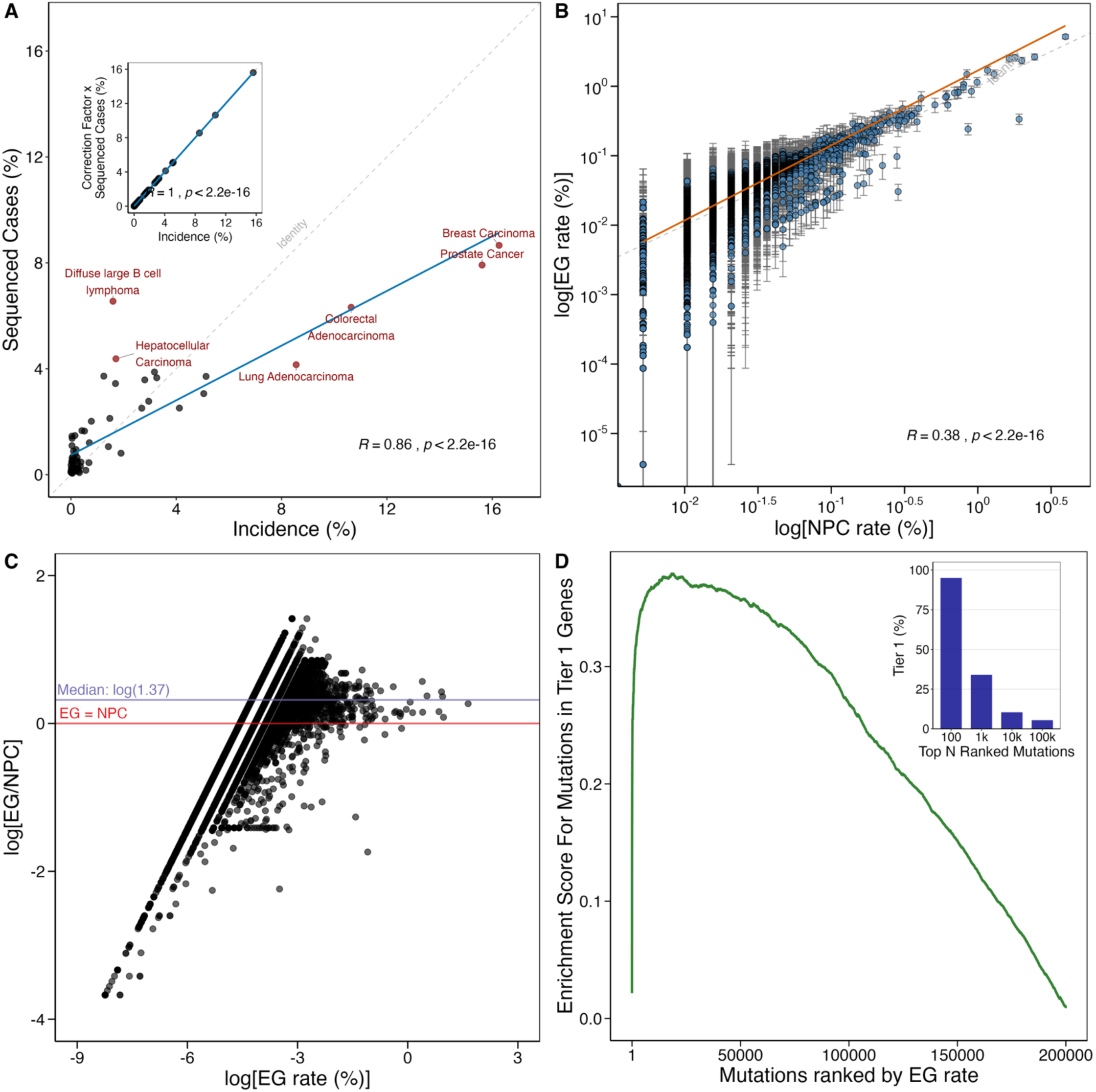
General characterization of the collection of missense and nonsense mutations. A. The proportion of all new cancer diagnoses attributed to the different forms of cancer (incidence %) and the proportion of all sequenced exomes considered that are for that same cancer type (sequenced cases %). The inset portrays the effect of applying the correction factor to the genomic data for the purpose of estimating specific missense and nonsense mutation rates within the US cancer patient population. B. For each missense and nonsense mutation, the log-transformed NPC rate and the log-transformed EG rate are plotted. The line of best fit for all missense and nonsense mutations is shown in red. C. For each missense and nonsense mutation observed more than once, the log-transformed EG rate and the log-transformed ratio of EG to NPC are plotted. The median EG to NPC ratio is 1.37. D. Enrichment plot for mutations in Tier 1 cancer driver genes among the set of all mutations observed more than once. The inset visualizes the fraction of mutations that occur in Tier 1 gene within the top 100, 1,000, 10,000, and 100,000 mutations ranked by EG rate.

We refer to the proportion of all exome samples that harbor a specific mutation as the “Naïve Pan-Cancer” (NPC) mutation frequency. This is consistent with conventional pan-cancer analyses that report mutation frequencies from the samples studied, and which have often been utilized as an estimate for frequencies in the general cancer patient population. We refer to the epidemiologically-incidence corrected frequencies as the “Epidemio-genomic” (EG) mutation frequencies.

The impact of epidemiological correction can be observed in several ways. For example, comparison of EG and NPC values finds a statistically significant trend that is offset from the identify, and where EG rates tend to be higher than the NPC rates (**Figure 1B**). Although there is a very good correlation, there is also significant dispersion away from the x=y axis (R=0.38, p<2.2e-16). Consideration of the fold change (i.e., EG/NPC) of mutation frequency corrections for all mutations finds a wide range of ratios between EG and NPC rates. This reveals median EG rates are 37% larger than their NPC counterparts.

EG values range from 97% larger than NPC rates (97.5% quantile) to 70% smaller than NPC rates (2.5% quantile) (**Figure 1C**). We additionally observe the most extreme values with the lower calculated EG Rates associated with only one or two observations in the data set. However, at higher calculated EG rates the range of EG to NPC ratios was much more narrowly distributed, highlighting reduced uncertainty for epidemiological corrections of the more recurrent mutations (i.e., those that are more commonly observed in cancer.)

Our set of cancer exomes harbors 2.11 million different mis-/nonsense mutations. Of these, 100,916 (or 4.8%) occur within a Tier 1 member of the Cancer Gene Census, which is a catalogue of known cancer drivers with multiple forms of evidence (*30*).

Consistent with the idea that cancer driver genes are selected for and are relatively more abundant, we find that the most common mis-/nonsense mutations are those that occur in known cancer driver genes. Specifically, 95 of the 100 most common mutations occur in a Tier 1 cancer driver (as well as 340 of the 1000 most common mutations) with the proportion progressively decreasing amongst the 10,000 most common mutations and 100,000 most common mutations (**Figure 1D – inset**). Across all 2.11 million mis-/nonsense mutations, we find that mutations occurring within any Tier 1 driver gene are statistically enriched (p-value < 0.001) (**Figure 1D**). Tier 2 genes of the Cancer Gene Census (p-value = 0.034) and genes that were neither Tier 1 nor Tier 2 members of the Cancer Gene Census (p-value 0.032) displayed no statistical enrichment (**Supplementary Figure 2**). These trends in enrichment are broadly consistent with principles of cancer biology and contribute to the validation of our bias correction methods.

### The most common missense and nonsense mutations

We next considered the most common missense and nonsense mutations in human cancer (**Supplementary Table I**). The raw, unbiased, data would provide inaccurate estimates for the most commonly observed mutations (**Supplementary Figure 3A**), justifying the need for our bias correction methods. Consideration of the twenty-five most prevalent missense and nonsense mutations finds ten mutations that are projected to be in at least 1% of the newly incident cases of malignant cancer (**Figure 3**). These ten most common mutations are found in four genes: *BRAF* (the single most common mutation), *KRAS* (second, fifth, eighth, and tenth most common), *PIK3CA* (third, fourth, and sixth most common), and *TP53* (seventh and ninth most common). *BRAF* V600E is the most common mutation within the incident cancer cases, being estimated to occur in 5.2% of all new cancer diagnoses. This is nearly twice as large as the second most common mutation, *KRAS* G12D, which is estimated to occur in 2.6% of all new cancer diagnoses. In addition to *KRAS*, *BRAF*, *PIK3CA*, and *TP53*, the twenty-five most common mutations also include mutations in *NRAS*, *AKT1*, *APC*, and *PGM5* (**Table I**).

**Figure 2.**
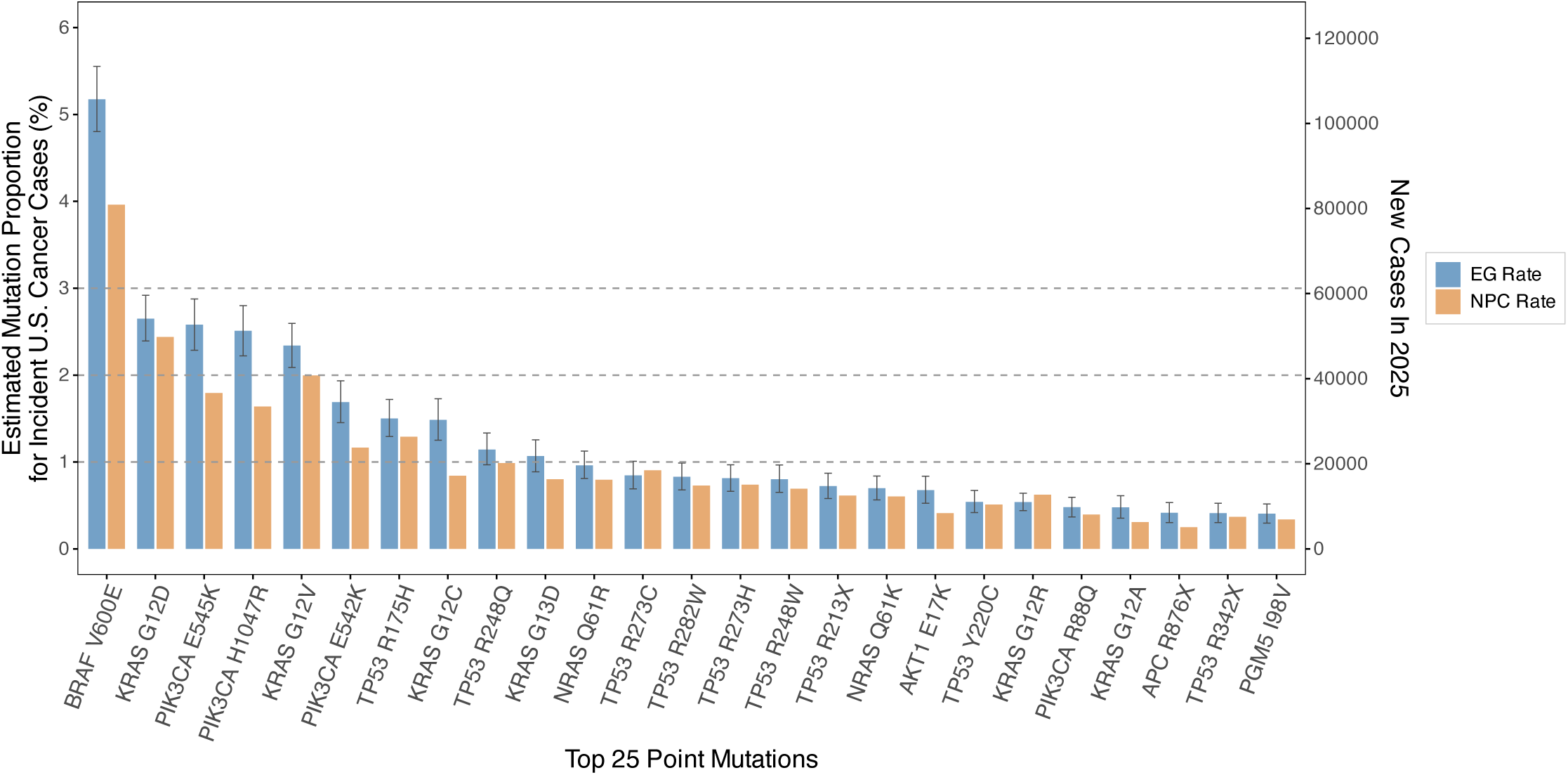
Calculated cancer patient mutation proportions (EG rates) for the most common missense and nonsense mutations. Estimated mutation proportion for incident cases in the US population and number of new cases in 2025 for the twenty-five most common missense and nonsense mutations ranked by EG rate.

**Figure 3.**
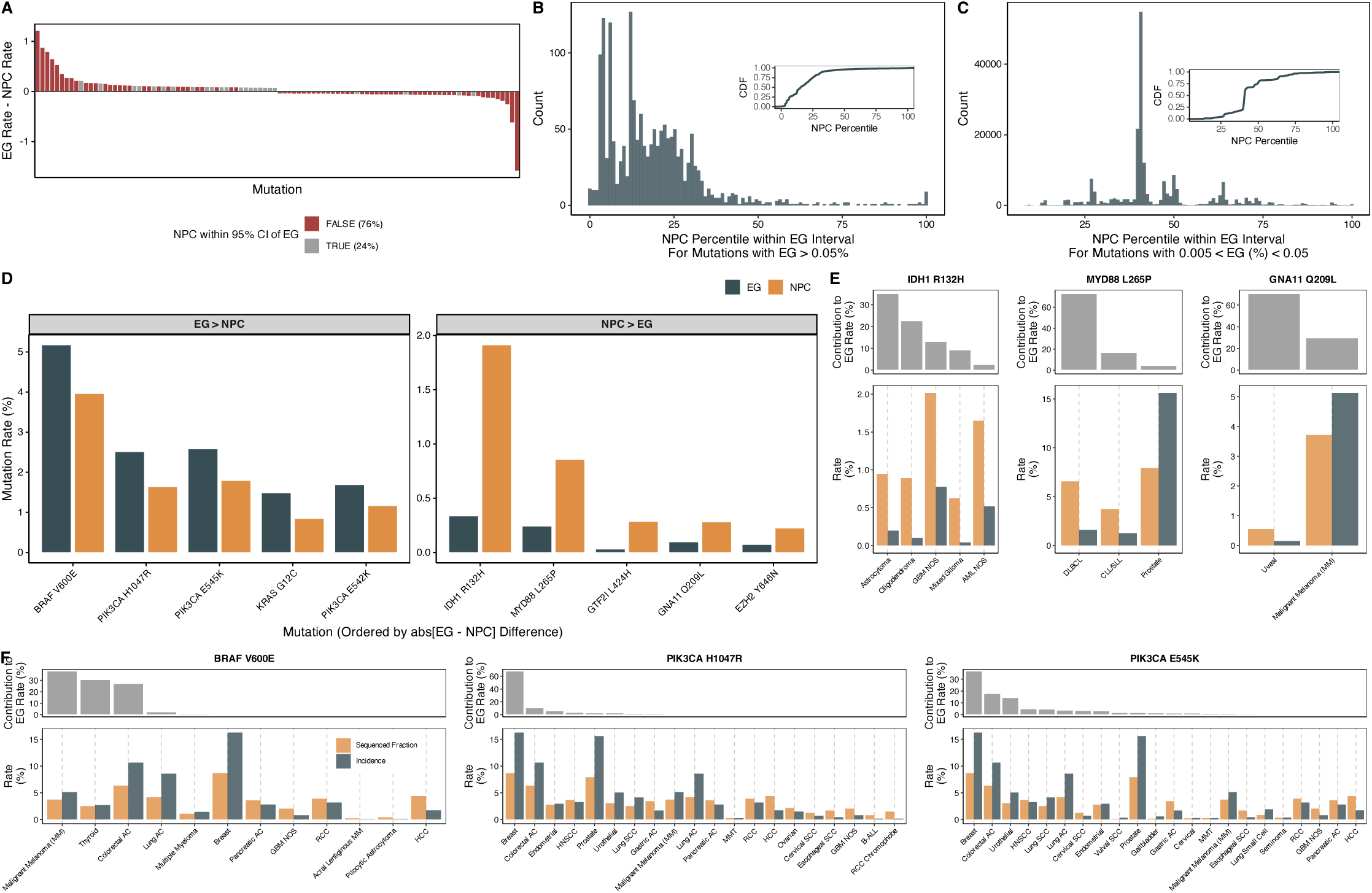
Evaluation of the discrepancies between population-level mutation proportions (epidemio-genomic, EG, rate) and the proportion of samples with the given mutation (naïve pan cancer, NPC, proportion). A. Waterfall plot of fifty largest EG-NPC difference mutations and fifty smallest EG-NPC difference mutations labelled by whether the NPC rate estimate falls within the 95% confidence interval of the EG estimate. B. Histogram of NPC percentile within the EG bootstrapped rate estimates for all mutations with EG rate above 0.05% and cumulative density function visualized in the inset. C. Histogram of NPC percentile within the EG bootstrapped rate estimates for all mutations with EG rate below 0.05% and cumulative density function visualized in the inset. D. EG and NPC estimates for the mutations with the largest magnitude differences between EG and NPC values. E. For *BRAF* V600E, *PIK3CA* H1047R, *PIK3CA* E545K, *IDH1* R132H, *MYD88* L265P, and *GNA11* Q209L, the contribution of that cancer type to the overall EG abundance for that mutation (grey, top subpanel) and the incidence (dark grey, bottom subpanel) and fraction of total sequenced cases belonging to the cancer type (orange, bottom subpanel). Cancer types shown represent 90+% of the mutations observed for the specified variant.

**Table 1.**
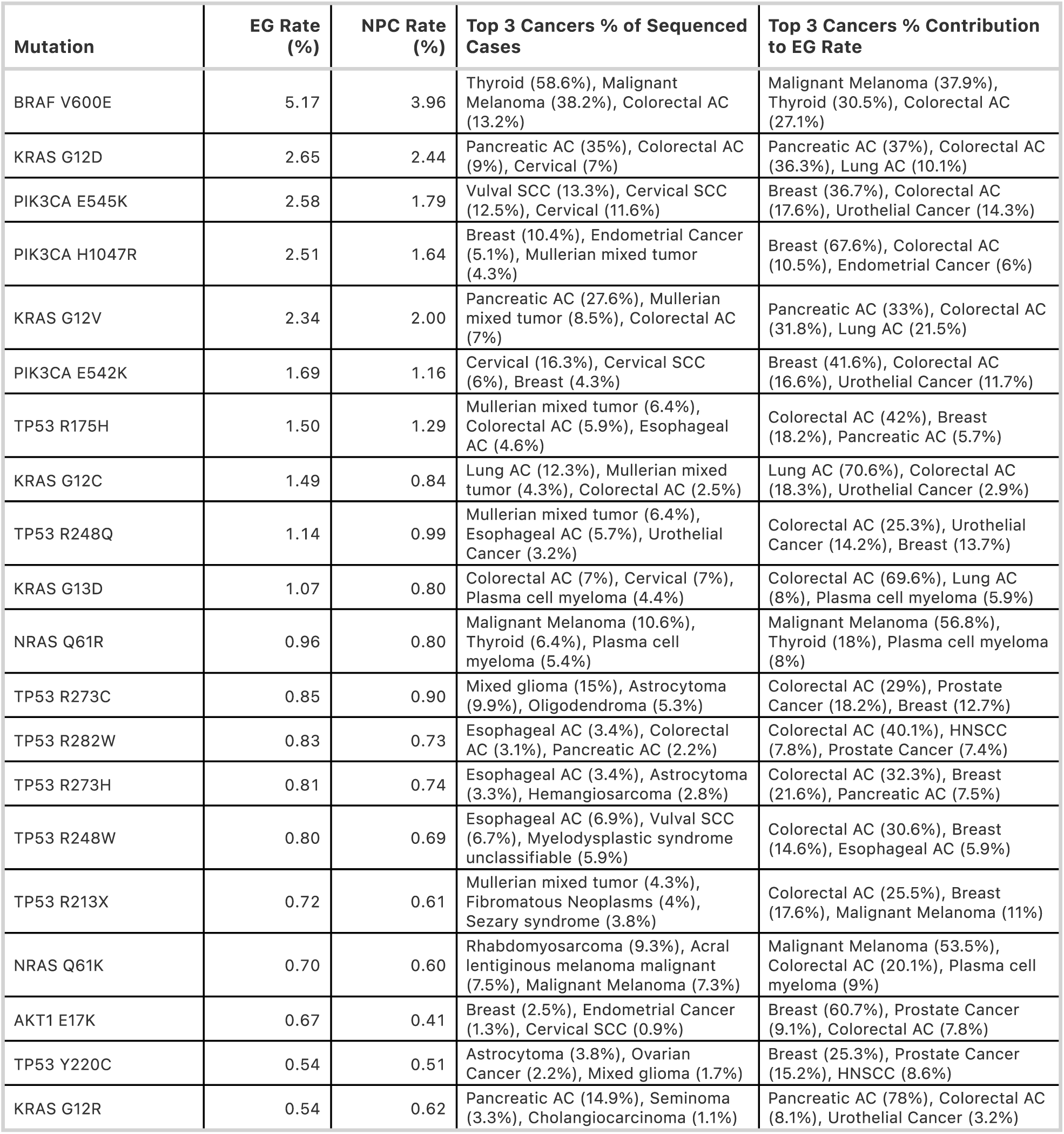
The twenty-five most abundant missense and nonsense mutations. For each of the twenty-five most abundant missense and nonsense mutations within the new cancer patient population (i.e., the highest epidemio-genomic (EG) rate), the specific mutation, its EG rate, its naïve pan cancer rate (NPC rate), the three individual cancers where this mutation is most abundant (and % of those cancer samples sequenced that harbor this specific mutation), and the three cancers that contribute the most to the overall EG rate (and the proportion of the final EG rate that this cancer contributes).

Consideration of 95% confidence intervals reveals high confidence in the projection that *BRAF* V600E is the most abundant mutation. *PIK3CA* E545K and *PIK3CA* H1047R are projected to be the third and fourth most common, at 2.6% and 2.5%, respectively. The only other missense mutation estimated to be found in more than 2% of all new cancer diagnoses is *KRAS* G12V, which is estimated to be found in 2.3% of all new diagnoses.

Consideration of the 95% confidence intervals reveals less certainty in the relative ordering of *KRAS* G12D, *PIK3CA* E545K, *PIK3CA* H1047R and *KRAS* G12V, but high confidence that these four mutations, in some ordering, represent the second through fifth most common acquired mutations in human cancer. Consideration of the 95% confidence intervals highlights that the epidemiologically-corrected incidence rates are commonly largely different from their non-weighted frequency in the cohort. For example, eight of the ten most common mutations have NPC rates that are outside of the EG 95% confidence intervals.

Consideration of the most common mis-/nonsense mutations that are in Tier 2 of the Cancer Gene Census highlights that specific mis-/nonsense mutations in genes with more limited and/or incomplete evidence for being a cancer driver are much less commonly encountered than the Tier 1 genes (**Supplementary Figure 3B, Supplementary Table I**). This does not necessarily indicate that these mis-/nonsense mutations do not play a role in cancer; it may rather be more difficult to establish driver roles for less commonly encountered mutations. Consideration of the most common mutations not in Tier 1 or Tier 2 of the cancer gene census is notable as the two most common mutations that are in neither Tier 1 or Tier 2 are actually more common than the most common Tier 2 mutations (*PGM5* I98V and *TRIM48* Y192H) (**Supplementary Figure 3C**). Notably, both genes have evidence for behaving like a tumor suppressor despite not currently being included within the Cancer Gene Census (*31–34*).

### Evaluation of the magnitudes of bias correction and their source

The overall magnitude of change introduced by implementing bias correction includes both large positive and large negative changes (**Figure 3A, Supplementary Table 1**). Notably, when we consider the fifty mutations with the largest positive absolute EG correction and the fifty mutations with the largest negative absolute EG correction, we find that 76% of these mutations have NPC rates that fall outside of the 95% confidence interval of the EG value, highlighting that NPC values are poor estimates for actual EG values.

Another approach to quantify the difference between EG and NPC rate estimates is to examine which percentile of the EG rate confidence interval distribution that the NPC estimate falls under. Mutations with an EG rate above 0.05% (n = 1662), have an NPC value that lies closer to the lower end of the EG rate estimate (**Figure 3B**). The median NPC value is at the 15^th^ percentile of the EG rate distribution. Among the fifty most common mutations, only five have an EG that is smaller than the NPC value (*TP53* R273C, 0.85 EG vs 0.90 NPC; *KRAS* G12R, 0.54 EG vs 0.62 NPC; *IDH1* R132H, 0.34 EG vs 1.91 NPC; *SF3B1* K700E 0.31 EG vs 0.32 NPC, *IDH1* R132C 0.31 EG vs 0.38 NPC). The strong trend for the more abundant mutations being underestimated in the biased set of cancer exomes is reduced for less common mutations: for mutations that occur less frequently, defined as an EG rate between 0.005% and 0.05% (n = 192,874), the median NPC estimate is at the 41^st^ percentile of the EG rate distribution (**Figure 3C**). Altogether, this suggests that less frequently encountered mutations tend to have concordant EG and NPC rate estimates whereas more frequently encountered mutations do not.

Consideration of the mis-/nonsense mutations with an EG level that is most underestimated by the NPC finds many mutations impacting important cancer driver genes, like *BRAF*, *PIK3CA*, and *KRAS* (**Figure 3D).** Consideration of which specific cancers contribute to their overall EG abundance highlights that these mis-/nonsense mutations tend to be most common in cancer types that have been disproportionately under sequenced relative to their overall population-level incidence (**Figure 3E**). For example, *BRAF* V600E has the largest positive correction (5.17% EG vs 3.96% NPC). Underscoring this discrepancy, the three cancers that contribute the most cases of *BRAF* V600E (malignant melanoma, thyroid carcinoma, colorectal adenocarcinoma) have all been disproportionately underrepresented in the cancer exome data. As another example, *PIK3CA* H1047R has the second largest positive correction the (2.51% EG vs 1.64% NPC), and the *PIK3CA* H1047R mutation has been underestimated in the overall cancer patient population because breast cancer, which contributes the majority of cases for this specific mutation, is approximately 50% underrepresented in the cancer exome data. Similarly, consideration of the missense mutations with an EG level that is most overestimated by NPC (**Figure 3D, right**) also finds many mutations in important cancer driver genes, like *IDH1* R132H (the mutation with the largest net change, with a 0.34% EG rate but a 1.91% NPC rate), *MYD88* L265P (the mutation with the second largest net reduction, with a 0.24% EG rate and a 0.86% NPC rate), and *GNA11* Q209L (the mutation with the fourth largest net reduction, with 0.10% EG rate and a 0.28% NPC rate) (**Figure 3E**). We find that the specific cancers that most contribute to these overestimated missense mutations tend to have been disproportionately over-sequenced relative to their population level incidence (**Figure 3E**).

### Gene level analysis of specific missense mutation relative abundances

A single driver gene will typically have multiple pathogenic alleles. Thus, it can be helpful to consider the relative proportions of the different mutations for a given driver gene. For example, the *KRAS* oncogene is important to cancer biology and to cancer pharmacology. The development of missense-specific targeting drugs for *KRAS* (28, 29) has generated more interest in understanding the relative abundances of the different *KRAS* mutations. We considered the EG and NPC rates for the different, observed, *KRAS* missense mutations and observe a strong linear correlation (R=0.99) with the best fit line off of the EG = NPC identify diagonal with high statistical significance, (**Supplementary Figure 4A**). *KRAS*, *NRAS*, *HRAS* are notable for having highly homologous amino acid sequences and similar structures (*35*), as well as for having very similar biochemical reaction rate constants (*36, 37*). It therefore seems likely that many of the missense-specific targeting drugs will be able to target mutations in *KRAS*, *NRAS*, and *HRAS* – and previous work has suggested that this is the case (*38, 39*). Our comparisons of NPC and EG rates for *NRAS* and *HRAS* find the same trends as for *KRAS*, with strong statistical significance for a high correlation (0.99 and 0.98) and with the best fits off of the EG=NPC identity diagonal with high significance (**Supplementary Figure 4B,C)**. This highlights the bias in the cancer genomic data and the need for epidemiogenomic corrections when estimating the abundance of specific (and potentially targetable) *KRAS*, *NRAS*, and *HRAS* missense mutations.

Our EG analysis finds the most common *KRAS* missense mutations are *KRAS* G12D, *KRAS* G12V, and *KRAS* G12C at 2.6%, 2.3%, 1.5% of all new cancer diagnoses, and projecting to approximately 54,000, 48,000, and 30,000 new patients diagnosed with a cancer containing these mutations per year (**Figure 4A**). Completely different missense mutations are most abundant for *NRAS* (**Figure 4B)** and *HRAS* (**Figure 4C**). Specifically, Q61R and Q61K are the first and second most common missense mutations for both *NRAS* and *HRAS*, with Q61L the third most common for *NRAS* and G12S the third most common for *HRAS*.

**Figure 4.**
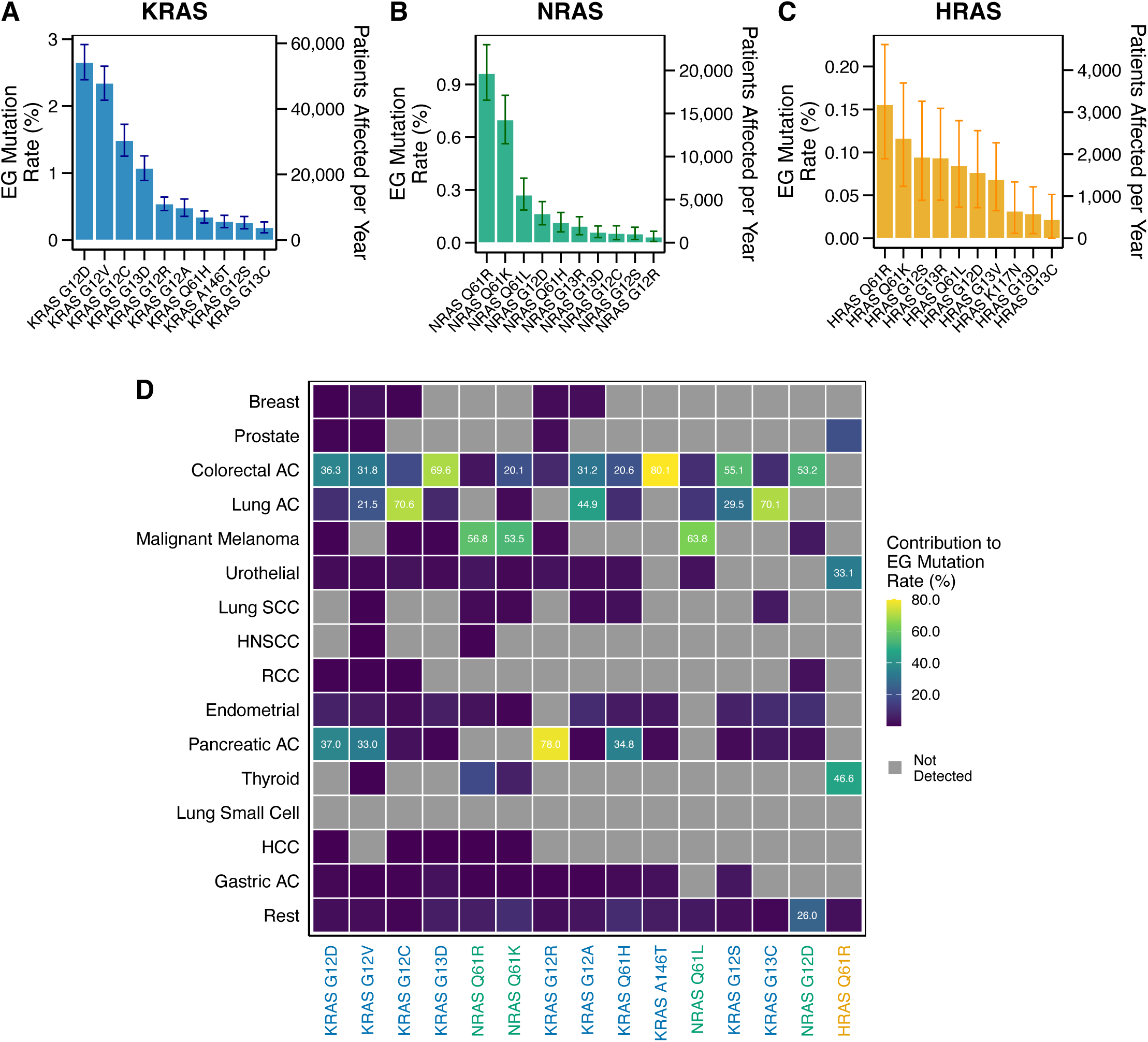
Evaluation of the most prevalent *KRAS*, *NRAS*, and *HRAS* mutations. A. Population level mutation abundance (EG rate) for the ten projected most common *KRAS* mutations, with 95% confidence intervals. B. Population level mutation abundance (EG rate) for the ten projected most common *NRAS* mutations, with 95% confidence intervals. C. Population level mutation abundance (EG rate) for the ten projected most common *HRAS* mutations, with 95% confidence intervals. D. Heatmap of the fifteen most prevalent *K/N/H-RAS* mutations visualizing the proportion of the overall EG rate that comes from each of the fifteen most common forms of cancer. When the proportion exceeds 20%, the percentage is displayed on the heatmap.

These data on *KRAS*, *NRAS*, and *HRAS* mutations can also be evaluated to determine which specific cancer types contribute the most cases of specific missense mutations (**Figure 4D).** This can be particularly informative when there are missense-specific pharmaceuticals (*28, 40*) or immunotherapies (*41*). Notably, *KRAS* A146T (*42*) is the missense mutation with the highest association with a single form of cancer: approximately 80% of all observations of *KRAS* A146T are found in colorectal adenocarcinoma. At nearly the same level of specificity is *KRAS* G12R (*43, 44*) for which 78% of new cancer diagnoses harboring this mutation are pancreatic adenocarcinoma.

The next highest associations include *KRAS* G12C (28, 29) and *KRAS* G13C (*45*) – with lung adenocarcinoma accounting for a little more than 70% of all observations for each mutation. Additionally, *KRAS* G13D (*26, 46–48*) displays high association with colorectal cancer, which accounts for nearly 70% of all observations. The most recurrent *NRAS* mutations, Q61R and Q61K, both have more than 50% of their occurrences accounted for by malignant melanoma. The most recurrent *HRAS* mutation, *HRAS* Q61R, is found most commonly in thyroid carcinoma (47%) and urothelial carcinoma (33%). Of the fifteen most common *RAS* gene mutations, ten had the majority of their cases coming from one specific form of cancer. The two most common *KRAS* mutations, *KRAS* G12D and *KRAS* G12V, were both less specific, with 37% and 36% of *KRAS* G12D cases coming from pancreatic adenocarcinoma and colorectal adenocarcinoma, respectively, and with 33%, 32%, and 22% of *KRAS* G12V cases coming from pancreatic adenocarcinoma, colorectal adenocarcinoma, and lung adenocarcinoma, respectively.

Another important cancer driver gene with many different recurrent mis-/nonsense mutations observed across many different forms of cancer is *TP53*. As with *KRAS*, the Pearson’s correlation between NPC rates and EG rates is 0.99 with a best fit line that is off of the diagonal with high statistical significance (**Supplementary Figure 4D**).

Consideration of the fifty most common *TP53* mis-/nonsense mutations finds a long tail of increasingly less common mis-/nonsense mutations (**Figure 5A**). All fifty of these mutations have been classified as “pathogenic” in at least one tumor context by the Universal Mutation Database (UMD) *TP53* Mutation Database (*49*), highlighting that all of these variants are cancer drivers with meaningful biological impact. The six most abundant individual missense mutations occur at four well-known hotspots: codons 175, 248, 273, and 282. These hotspots fall within the DNA-binding domain (DBD) of *TP53* (*50*), which spans from codon 94 to codon 292 (*51*). Notably, only six of the projected top fifty mutations do not occur within the DBD: *TP53* R342X (9^th^ most common), R306X (12^th^), Q331X (23^rd^), E298X (30^th^), Q317X (31^st^), E294X (36^th^).

**Figure 5.**
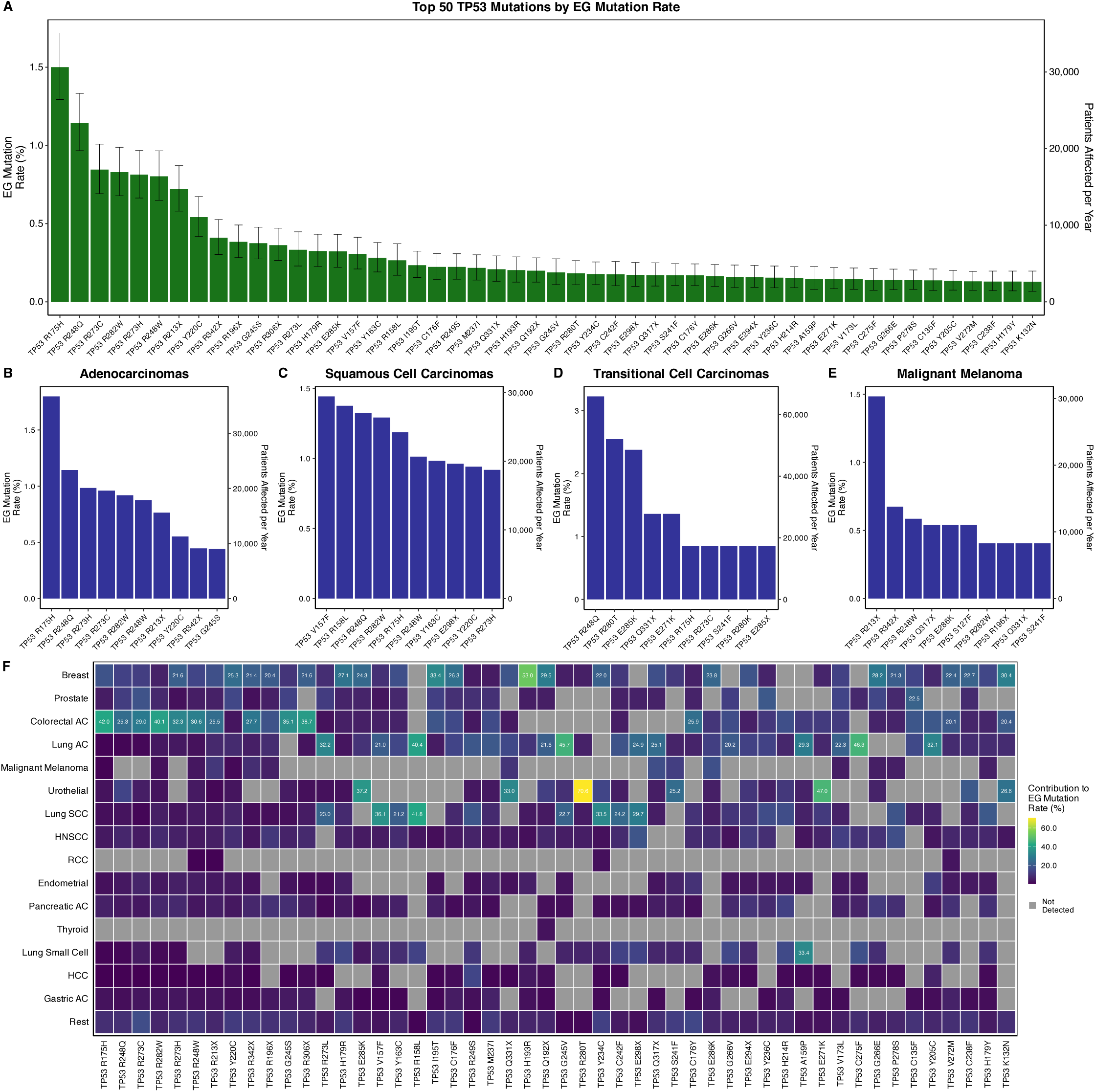
Evaluation of the most prevalent *TP53* mutations. A. Population level mutation abundance (EG rate) for the fifty projected most common *TP53* mutations, with 95% confidence intervals. B. Population level mutation abundances (EG rate) for the ten most common TP53 mutations within the subset of the cancer patient population with a form of adenocarcinoma. C. Population level mutation abundances (EG rate) for the ten most common TP53 mutations within the subset of the cancer patient population with a form of malignant squamous cell carcinoma. D. Population level mutation abundances (EG rate) for the ten most common TP53 mutations within the subset of the cancer patient population with a transitional cell carcinoma. E. Population level mutation abundances (EG rate) for the ten most common TP53 mutations within the subset of the cancer patient population with a form of melanoma. F. Heatmap of the fifty most prevalent *TP53* mutations visualizing the proportion of the overall EG rate that comes from each of the fifteen most common forms of cancer. When the proportion exceeds 20%, the percentage is displayed on the heatmap.

Our previous, gene-level, EG analysis found that not only was *TP53* the most commonly mutated gene in all of cancer, being found in an estimated 35% of all newly diagnosed cancers in the US (19), it was also the most common in three major classes of cancer: adenocarcinoma (34%), squamous cell carcinoma (63%), and transitional cell carcinoma (44%). It was the 18^th^ most commonly mutated gene in malignant melanomas, the fourth cancer class evaluated, at 15% of samples mutated. We evaluated the most common mis-/nonsense *TP53* mutations in these four major classes of cancer and observed markedly different patterns of occurrence. For example, *TP53* R175H, which is the most common *TP53* mutation across all cancers, is also the most common *TP53* missense mutation when the analysis is limited to adenocarcinomas (**Figure 5B**). However, *TP53* R175H is only the 5^th^ most common missense mutation when the analysis is limited to malignant squamous cell carcinomas (**Figure 5C**) and the 6^th^ most common when the analysis is limited to transitional cell carcinomas (**Figure 5D**). Notably, *TP53* R175H is not even among the ten most common *TP53* mutations in malignant melanoma (**Figure 5E**); rather, *TP53* R213X is the most common *TP53* mutation in malignant melanoma. Nonsense mutations in *TP53* are particularly important for malignant melanoma; five of the ten most common mutations in *TP53* within malignant melanoma are nonsense mutations. In contrast, nonsense mutations make up only two of the ten most common *TP53* mutations in adenocarcinoma and transitional cell carcinoma, and only one of the ten most common *TP53* mutations in malignant squamous cell carcinomas.

As opposed to the *RAS* gene mutations, where it was common for an individual missense *RAS* mutation to have more than 50% of its observations across all cancers attributable to a single form of cancer, *TP53* missense mutations are less predominantly associated with any single form of cancer (**Figure 5F**). Only two of the fifty most prevalent *TP53* mutations had the majority of new cases due to a single cancer: urothelial carcinoma for *TP53* R280T (71%) and breast carcinoma for *TP53* H193R (53%). *TP53* R158L was almost exclusively associated with forms of lung cancer (42% lung squamous cell carcinoma, 40% lung adenocarcinoma, 14% lung small cell carcinoma). Although several other *TP53* mutations had their majority of cases associated with the three lung cancers (*TP53* 273L, *TP53* V157F, *TP53* G245V, *TP53* E298*) these mutations were observed in several other forms of cancer, in contrast to the more specific *TP53* R158L mutation. Associations with both lung adenocarcinoma and lung squamous cell carcinoma were not universal; for example, 34% of total *TP53* Y234C occurrences are projected to be found in lung squamous cell carcinomas but it is projected to rarely be observed in lung adenocarcinoma. Additionally, 46% of *TP53* C275F occurrences are projected to be found in lung adenocarcinomas and to be rarely observed in lung squamous cell carcinoma.

Evaluation of which tumor types routinely harbor specific *TP53* mutations finds that the ten most abundant *TP53* mutations are often found in the four most common forms of cancer: breast carcinoma, prostate carcinoma, colorectal adenocarcinoma, and lung adenocarcinoma (**Figure 5F**). Some tissue-specific associations of *TP53* with specific carcinogenic processes have previously been described (*52, 53*), however the interplay between carcinogenic processes and contextual selective factors in different tumors remains incompletely understood.

The data in Supplementary Table 1 include information on all recurrent missense and nonsense mutations, and Supplementary File 1 includes data on all missense and nonsense mutations. These tables are sufficient to enable analyses of other genes as was done here for *KRAS*, *NRAS*, *HRAS*, and *TP53.* This file also contains the total number of mutations observed across all cancers, mutation counts in each cancer type, EG rate, NPC rate, gene name, and specific mutation change (e.g., KRASp.G12C), thereby enabling analysis of any number of genes or gene sets.

### Gene level analysis of classes of missense mutations

It is well appreciated that many oncogenes have one or more “hot spots” where mutations tend to cluster. It is also well appreciated that the different oncogenic forms of a given proto-oncogene can vary with respect to observable behaviors like magnitude of response and sensitivity to targeted therapies (*54*). Our data enables analysis of hotspots and functional classes. We therefore utilized these data to analyze *RAS*, *BRAF*, and *TP53* mutations further by their respective, important, subclasses.

The major hotspots for the *RAS* genes are at codons 12, 13, and 61. Minor hotspots have been described at several codons, including 146, 117, and 59 (*55*). We evaluated the relative abundances of different hotspots for *KRAS* (**Figure 6A**), *NRAS* (**Figure 6B**), and *HRAS* (**Figure 6C**). The absolute EG rate estimates for each hotspot *H/N/KRAS* codon is reported in **Supplementary Figure 5A-C**.

**Figure 6.**
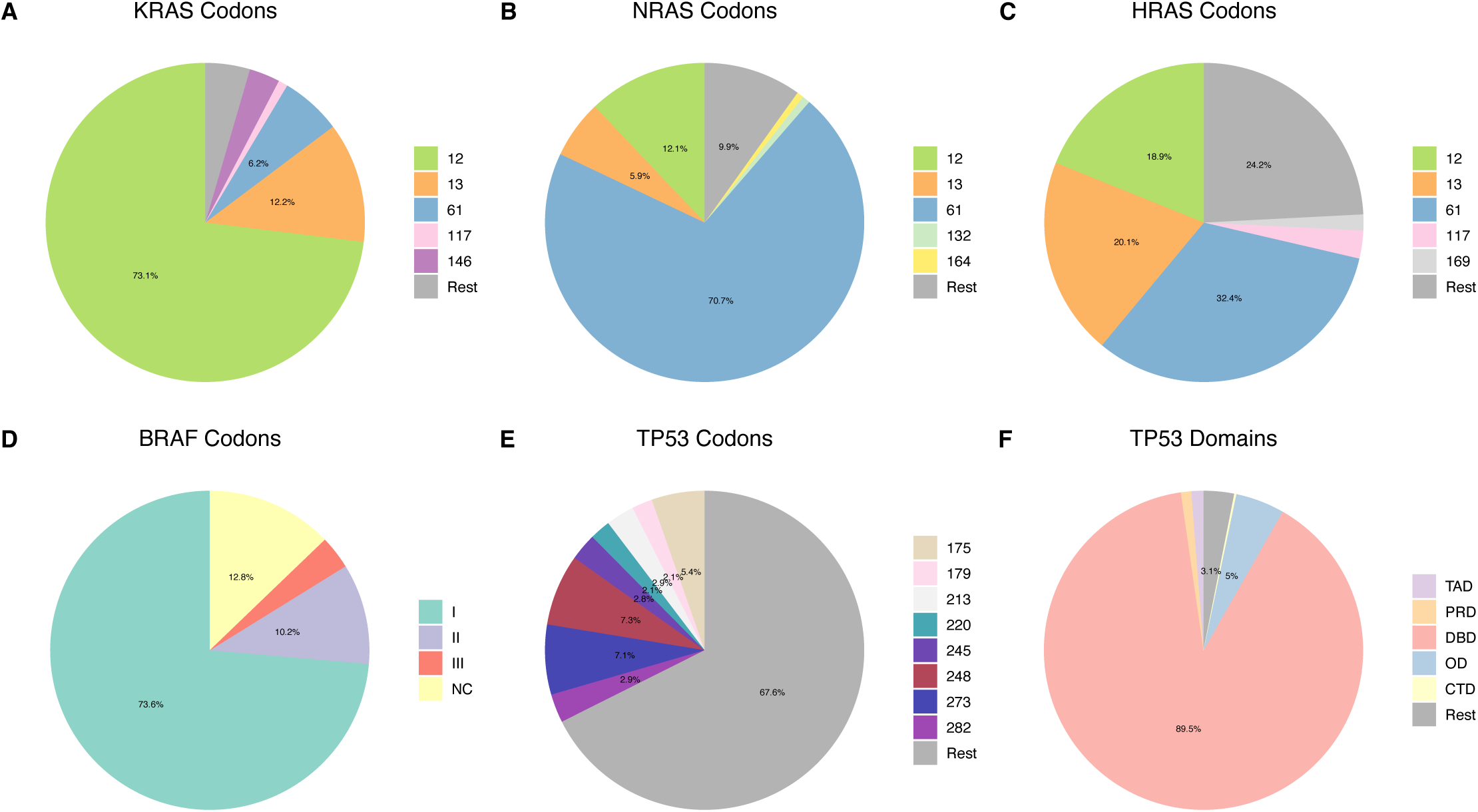
Analysis of important classes of *KRAS*, *NRAS*, *HRAS*, *BRAF*, and *TP53* mutations. A. Pie chart showing the proportion of *KRAS* mutations that occur in codon 12, 13, 61, 117, 146 (i.e., the five most commonly mutated *KRAS* codons), or any other codon. B. Pie chart showing the proportion of *NRAS* mutations that occur in codon 12, 13, 61, 132, 164 (i.e., the five most commonly mutated *NRAS* codons), or any other codon. C. Pie chart showing the proportion of *HRAS* mutations that occur in codon 12, 117, 61, 117, 169 (i.e., the five most commonly mutated *HRAS* codons), or any other codon. D. Pie chart showing the proportion of *BRAF* mutations that are best described as Class I, Class II, Class III, or not yet classified. E. Pie chart showing the proportion of TP53 mutations in codons that account for at least 2% of all TP53 mutations, or any other codon. F. Pie chart showing the proportion of TP53 mutations in each functional domain of TP53. TAD = Transactivation domain, PRD = Proline-rich domain, DBD = DNA-Binding Domain, OD = Oligomerization domain, CTD = Carboxy-terminal domain.

This analysis demonstrated that codon 12 accounts for nearly 75% of *KRAS* mutations in the U.S. Cancer Patient Population, while codon 12 only accounts for 12% of *NRAS* and 19% of *HRAS* mutations. Rather, codon 61 is the dominant hotspot mutation for *NRAS* (71%) and *HRAS* (33%). Notably, codons 12, 13, and 61 comprise 72% of the *HRAS* mutations and a long tail of less common mutations accounts for the remaining 28%, with only one minor hotspot, codon 117 (3%), projected to account for more than 2% of all observed *HRAS* mutations. Altogether, this analysis characterizes the important *RAS* oncogenes at the level of major and minor hotspots. Importantly, this analysis differs from previous characterizations of *RAS* that was done through analysis of biased cancer genome data sets (*56, 57*) and thus highlights the value of bias correction.

Additionally, we considered functional classes of mutations. For example, the *BRAF* oncogene has been divided into three classes: Class I, Class II, and Class III based on the behavior of the corresponding oncoprotein (*25*). Notably, Class I *BRAF* mutations result in a protein with an active conformation, even without dimerization (*25, 58*). Class II *BRAF* mutations result in a protein that achieves an active conformation in a dimer-dependent and RAS-independent manner, and Class III *BRAF* mutations activate wild-type RAF counterparts and hence turn on downstream signaling in a dimer-dependent and RAS-dependent manner (*25*). Many of the *BRAF* mutations observed in cancers have been studied at a level that enables characterization as Class I, II or III (*25, 58–63*). However, many observed *BRAF* mutant alleles remain uncharacterized.

We evaluated the EG relative and absolute abundances of Class I, II, and III *BRAF* mutations, as well as those yet to be characterized (**Figure 6D, Supplementary Figure 5D**). *BRAF* Class I, which includes *BRAF* V600E, the single most EG abundant mis-/nonsense mutation in the US cancer patient population, was found to comprise 74% of all observed BRAF mutations. Class II and Class III each comprised 6% of all observed *BRAF* mutations. The value for all three classes is likely to change as more *BRAF* mutations are characterized because 13% of all observed *BRAF* mutations have yet to be characterized. Notably, this analysis is limited to missense and nonsense mutations and not to *BRAF* fusion proteins (*64*). Additionally, although the *BRAF* mutations found in 13% of the cancer patient population that have yet to be classified may potentially fall into Class I, II, or III, it is also likely that some may be passenger mutations. Additionally, it is possible that one or more additional classes of *BRAF* mutations may ultimately be identified, especially considering the complex regulation of *BRAF* activation (*65*) and the non-intuitive consequences that can follow from perturbation of this pathway (*66–72*). We note that our proportions are different from a previous, large (>110,000 patients), pan-cancer analysis of Class I, II, and III BRAF mutations that found 62% of BRAF mutations were Class I, 17% were Class II and 18% were Class III (*73*); however, that cohort included many cancer types that were over sampled (and under sampled), relative to their population level proportion among incident cancer patients.

We evaluated *TP53* mutations both by codons and by domains (**Figure 6E,F**). *TP53* codons 248, 273, and 175 account for 7.3%, 7.1%, and 5.4% of all observed mutations, respectively (**Supplementary Figure 5E**). The finding that these three codons are the most commonly mutated is consistent with previous pan-cancer aggregations (*51*). However, our analysis provides enhanced accuracy with epidemiologically-based frequencies.

Mutations were observed in 220 total codons in our data set, with twenty-nine of them accounting for 1% or more of the total mutations (**Supplementary File 1**). 90% of *TP53* missense and nonsense mutations occur in the DNA Binding Domain (DBD) (**Figure 6F**). This corresponds to 27% of incident cancer cases harboring a mutation in the DBD of *TP53* (**Supplementary Figure 5F**).

### Pathway level analysis

Although there are many forms of cancer with different etiologies, shared biological phenotypes have been noted across all cancers (*74, 75*). For example, self-sufficiency in growth signals is an important hallmark of cancer. This phenotype is commonly imparted by a cancer driver mutation within the RTK/RAS signaling pathway. We investigated which missense and nonsense mutations were most common within the members of the RTK/RAS pathway using a previously defined component list for this pathway that was developed for evaluating cancer genomics data (*4*). The projected twenty-five most common missense and nonsense mutations in this pathway involves mutations to *BRAF* (two), *KRAS* (ten), *NRAS* (four), *FGFR3* (one), *ERBB2* (four), *EGFR* (one), *RAC1* (one), *HRAS* (one), and *ERBB3* (one) (**Figure 7A**). Thus, although mutations in *KRAS*/*NRAS*/*HRAS* comprise more than half of these mutations, an additional six different genes contribute important individual RTK/RAS pathway driver gene mutations.

**Figure 7.**
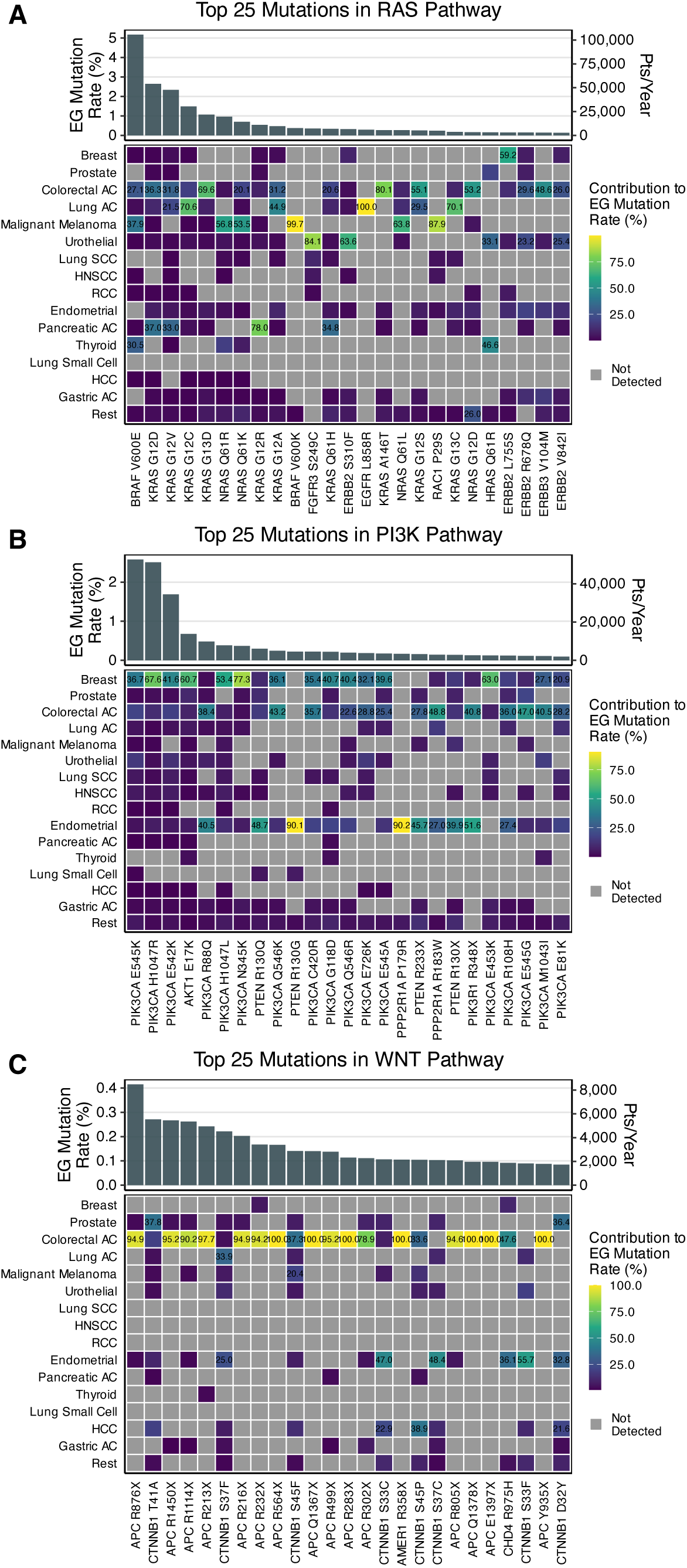
Analysis of the most common mutations in important cancer cell signaling pathways. (Top) Bar charts showing the EG rate and projected number of new malignant cancer diagnoses in the U.S. for each mutation for the corresponding pathway. (Bottom) Heatmaps with the proportion of the overall EG rate that comes from each of the 15 most common forms of cancer for the corresponding pathway. When the proportion exceeds 20%, the percentage is displayed on the heatmap. A. Population level mutation abundance (EG rate) for the twenty-five projected most common RAS/RTK pathway mutations, with 95% confidence intervals. B. Population level mutation abundance (EG rate) for the twenty-five projected most common PI3K pathway mutations, with 95% confidence intervals. C. Population level mutation abundance (EG rate) for the twenty-five projected most common WNT/β-catenin pathway mutations, with 95% confidence intervals.

In addition to providing the single most abundant individual cancer-associated mutation (overall, not simply within the RTK/RAS pathway), *BRAF* also provides the tenth most frequently encountered RTK/RAS pathway mutation (*BRAF* V600K). Following these two mutations, there is a steep fall off for the frequency with which *BRAF* mutations are observed, with the next most common falling to 28^th^ (*BRAF* K601E) and 32^nd^ (*BRAF* G469A) most common (**Supplementary Table 1**). The tissue specific patterns of these mutations demonstrate that *BRAF* V600K is projected to be almost exclusively found in melanoma patients. This is consistent with it being almost exclusively found in melanoma within other genomic databases that aggregate a wider range of genomic data (*30*). This is in contrast to the V600E mutation, which – at the cancer patient population level – is most commonly found in patients with melanoma (37% of all V600E incident cases), thyroid carcinoma (30% of all V600E incident cases), and colorectal adenocarcinoma (27% of all V600E incident cases). Other examples of high context specificity beyond *BRAF* V600K include *EGFR* L858R (90% of occurrences associated with lung adenocarcinoma), *RAC1* P29S (87%, melanoma) (*18*), *ERBB2* S252W (93%, endometrial), and *FGFR3* S249C (84%, urothelial).

We also considered the projected twenty-five most common mutations in the PI3K signaling pathway (**Figure 7B**). These were predominantly *PIK3CA* (seventeen of the twenty-five) but also included one *AKT1* mutation, *AKT1* E17K (*76*); four *PTEN* mutations (R130Q, R130G, R233/*, and R130/*), two *PPP2R1A* mutations (P179R and R183W) and one *PIK3R1* mutation (R348/*). Of these twenty-five mutations, only eight have the majority of their cases attributable to a single form of cancer. This is in contrast to the RAS pathway, where sixteen of the twenty-five most abundant mutations are attributable to one form of cancer. Additionally, the extent of specificity was less for the PI3K pathway, with only two mutations at 80% or above (*PTEN* R130G, 90%, endometrial; *PP2R1A* P179R, 90%, endometrial). The six most abundant PI3K pathway mutations were commonly found in more than half of the fifteen most commonly diagnosed forms of malignant cancer, while the remaining nineteen most common PI3K pathway mutations were generally found in a smaller number of cancer types.

In addition to the RTK/RAS and PI3K networks, which are commonly disrupted by oncogenes, we also considered the WNT/ β-catenin pathway that is commonly dysregulated by tumor suppressor mutations. We project fewer highly recurrent mutations in this pathway, in part reflecting the large number of possible ways for a tumor suppressor to be inactivated by a mutation that results in loss of function. Consideration of the projected twenty-five most abundant WNT/ β-catenin pathway mutations (**Figure 7C**) revealed that *APC* contributed the most (fifteen, with all of them being nonsense mutations), with *CTNNB1* contributing eight mutations, and *AMER1* and *CHD4* each contributing one. All of these specific mutations have evidence for pathogenicity (*77, 78*). The heat map showing which cancers contribute to the projected abundances of each mutation show a high contribution from colorectal cancer for nearly all of these mutations, consistent with the well-established role of this pathway in colorectal cancer tumorigenesis (79). Notably, each *APC* mutations had the majority of the projected population level incidence to come from colorectal cancer. Mutations in *CTNNB1* were much more widely distributed between cancers, with prostate cancer contributing the largest fraction of *CTNNB1* T41A mutations (the 2^nd^ most abundant in the WNT/ β-catenin pathway). Prostate cancer also contributes a plurality of cases *CTNNB1* D32Y (36%), with endometrial carcinoma also contributing a large fraction (32%). Endometrial cancer contributed the plurality to *CTTNB1* S33C (46%), *CTTNB1* S37C (48%), and *CTNNB1* S33F (55%). Colorectal adenocarcinoma, lung adenocarcinoma, malignant melanoma, and hepatocellular carcinoma were all also important contributors to one or more of the most abundant *CTNNB1* mutations. Relative to the RAS/RTK and PI3K pathways, the WNT/β-catenin pathway shows less evidence for being commonly mutated across the most common forms of malignant cancer.

Altogether, consideration of genes from specific pathways, complexes, or processes can identify which specific missense and/or nonsense mutations are most commonly contributing to the development and maintenance of cancer phenotypes. The set of recurrent mutations provided in Supplementary Table 1 enables similar analyses to be performed for the pathways, complexes, and processes of one’s choice.

## DISCUSSION

Altogether, we present a compendium of missense and nonsense mutations that have been observed in human cancer and that includes our projection for what percentage of the newly diagnosed cancer patient population harbors that specific missense or nonsense mutation. We also provide 95% confidence intervals for these projections to help qualify the data, information on which specific cancers were observed to harbor the mutations, the counts of observations for specific cancers, and the percentage of the incident cancer population that comes from each specific missense or nonsense mutation in each cancer (**Supplementary File 1**). From these data, the analyses demonstrated in this study could be performed for other genes and pathways beyond the presented demonstrative examples that focus on several high priority genes and pathways.

We envision that these data should be broadly useful. For example, cancer researchers have commonly utilized various types of pan-cancer analyses and databases to estimate how commonly a given gene is mutated in cancer. Although pan-cancer studies offer many values, the percentage of samples with a given mutation from a collection of cancer samples that is not strictly proportional to the incidences of those different cancers will generally not equal the actual percentage of cancer patients in the population with that mutation, and may rather give a poor estimate. Additionally, many published pan-cancer analyses are limited to a small fraction of the many different forms of cancer. Our data set includes samples from cancer types that are representative to 93% of the diagnoses within the U.S. cancer patient population.

Examples of differences between our epidemiologically-informed genomics-based estimates for mutation prevalences and values provided in the literature from pan-cancer analyses include *KRAS*, *NRAS*, and *HRAS* codon breakdowns (*56, 57*), *KRAS*, *NRAS*, and *HRAS* allele breakdowns (*80–82*), *BRAF* Class I/II/III breakdowns (*83*), and *TP53* mutant allele proportions and codon proportions (*84*). Although trends were often similar, values and relative orderings typically diverged which further highlights that epidemiological correction impacts understandings of genomic data.

A better understanding of the relative abundances of different mutations across the cancer patient population can also be useful for basic science and translational cancer research. For example, many cancer researchers have developed and/or utilized isogenic series model systems in which one model system exists in different forms with each form harboring a different pathogenic mutant allele (*26, 27, 48, 85–88*). When focusing exclusively on a single cancer type, the previously existing genomic data have served well for identifying the most important mutations for that specific form of cancer being studied. However, with our corrected data, researchers can now better consider how common mutations from their particular cancer of interest also fall within the broader cancer patient population. As researchers typically focus on a subset of the available mutations, they may choose between mutations that are similarly abundant in their cancer type of interest based on how abundant they are in the broader population to have the chance to make their research even more impactful.

Another value for these data is in the evaluation of drug targets. One emerging class of targetable missense mutations has been acquired cysteine residues. For example, KRAZATI and LUMAKRAS are small molecule inhibitors that covalently react with the acquired, nucleophilic, cysteine residue at codon 12 of *KRAS* G12C (*28, 29*). The success of these compounds has led to interest in an expansion of the covalent targeting approach to other acquired nucleophiles (*89, 90*). We have previously presented a catalogue of acquired cysteine residues that communicates the population level abundance of different acquired cysteines in the cancer patient population (*22*). The data included in Supplementary Table I can be analyzed to identify the acquired nucleophiles that are projected to be most abundant. This analysis would identify the nucleophilic drug targets that are most common in the cancer patient population.

Although this study corrects for cancer-type bias in pan-cancer analyses, many other forms of bias exist in cancer genomics. For example, non-proportionate representation of individuals between sexes, age groups, and genetic ancestry groups has previously been described to be a problem within cancer genomics (*91*). Additional variables that can also bias the results of cancer genomics analyses include the stage of the tumor, how soon after initial diagnosis the tumor was sampled, and the extent of treatment that the patient has received prior to sequencing. Next steps in the extension of bias correction methods in cancer include correcting for additional forms of bias. Because of the various potential areas for bias, all cancer genomic data should be considered an approximation of the underlying real-world system of interest.

Although bias correction methods should result in enhanced estimation accuracy, some extent of uncertainty in the estimates will remain. We provide 95% confidence intervals as part of our analysis. The magnitudes of these ranges highlight that the existing data are sufficient to generate good estimates for population level abundances of specific missense mutations, but that relative ordering of mutations with similar projections may differ in the patient population. This uncertainty follows from individual mutations being relatively uncommon, as only five mutations are projected to be found in more than 2.5% of the cancer patient population.

Another source of uncertainty follows from gaps in the data. For example, the available exome data include samples that represent approximately 93% of all cancer diagnoses. For the 7% that do not have representative exomes, we assume the overall weighted prevalences from the 93% with data are reasonable approximations. This is a common method for imputing missing data within data science studies. These 7% of cancers represent hundreds of different rare cancer types. Some rare cancer types always/almost always have a very specific mutation; for example, almost all classic hairy cell leukemia cases harbor a *BRAF* V600E mutation (*92*). However, this cancer type is also rare, accounting for 0.06% of all new cancer diagnoses (*9*), and therefore account for less than 1% of the cancers for which we are imputing values. As a large fraction of cancer types have no/rare levels of *BRAF* V600E mutations, the imputation approach seems reasonable for the most commonly encountered mutations. However, for most of the 200,000 specific recurrent mutations that are part of this study, a rare cancer as common as hairy cell leukemia with a single mutation that appears in nearly all cases will make a contribution that exceeds the imputed value and the 95% confidence intervals. Nevertheless, we anticipate that the number of pathognomonic mutations that exceed the imputed value (and possibly the 95% confidence interval limits) will be a very small fraction of the more than 200,000 missense mutations analyzed in our study.

Altogether, this analysis presents estimates for the abundance of specific missense and nonsense mutations within the cancer patient population. These data present a useful portrayal of cancer genetics that will be useful for a variety of basic science, translational, and clinical research applications. We also anticipate that this approach of correcting for biases in the genomic data to obtain more accurate portrayals of specific populations will continue to develop as a useful analytical approach.

## METHODS

### Collection of Genomic and Epidemiological Data

Cancer epidemiological data were obtained from the NCI Surveillance Research Program database SEER using SEER*Stat software (*9*). We utilized the database named Incidence – SEER Research Data, 18 Registries, Nov 2019 Sub (2000–2017). We exported data in table output with rows specified by ICD-O-3 Histo/behave codes and columns specified by Site recode ICD-O-3/WHO 2008 location codes. To obtain epidemiologically-informed estimates for mutation rates, we assign a ROSETTA code to each SEER ICD-O-3 code. As previously described, the ROSETTA code is a unified set of codes that aligns cancer type descriptions between SEER and cBioPortal data (*19*). We collect data from 140 different publicly available cancer exome studies sourced from cBioPortal (*5*) and assign a ROSETTA classification for each individual tumor sample based on the metadata associated with that tumor sample. If a patient appears multiple times across studies, we keep the first collected sample. Mutations with no assigned HUGO symbol are removed.

We have representative sequencing data from cBioPortal for ROSETTA codes that accounted for 93% of all observed human cancers. Thus, we normalized the SEER incidence vector “R” subset to the codes that accounted for these 93% of cases to arrive at the standard incidence vector “I” (Supplementary Figure 1). We did not acquire tumor samples from the 7% of cancer diagnoses not well characterized by whole exome studies and assume that the weighted proportions of mutations for these 93% of all cancer are a good estimate for the weighted proportion in all cancers (*19*).

### Estimation of the epidemiogenomic (EG) mutation rate and confidence intervals

We assemble a count matrix where each entry represents the number of observations of a given mutation in a specific cancer type represented as matrix “C” in Supplementary Figure 1. Then, we compute a vector of incidence fractions “I” for each cancer type, as described above. Finally, we compute a separate vector “N” with the number of total sequenced tumors for each cancer type. For a given mutation (i.e., row in count matrix), we compute the element-wise ratio between the incidence and sequenced cases vector and multiply it by the mutation observations in that cancer type. Summing these values across cancer types results in an epidemiogenomic (EG) estimate for a given mutation (Supplementary Figure 1). To estimate the number of incident cases at the mutation level, we multiply the EG rate estimate by 2,041,910, the number of new malignant cases estimated to occur in 2025 in the U.S. (*93*).

Confidence intervals for each mutation were obtained by a bootstrap approach. We created 10,000 re-sampled count matrices. For each specific mutation within each specific form of cancer, we assumed that the observed number of patients, amongst those sequenced, with the mutation in question was well-described by a Poisson distribution. For each entry in the re-sampled count matrix (i.e., for each specific mutation within each specific form of cancer), we drew a random sample from a Poisson distribution with parameter equal to the value of that entry in the original “C” count matrix. This process was repeated 10,000 times. Each re-sampled count matrices were then multiplied by the Incidence vector to obtain 10,000 sets of re-sampled EG estimates for each mutation. The 2.5% and 97.5% quantile from the re-sampled EG rate estimates were used to construct 95% confidence intervals for each mutation (Supplementary Figure 1).

The naïve pan-cancer (NPC) mutation rate for a given mutation may lie within the bounds of the 10,000 EG bootstrapped rates. In these cases, the percentile in the EG distribution that the NPC rate occupies is computed (via the percentileofscore function in python). If it is greater than all the bootstrapped rates, the NPC rate is noted to lie in the 100^th^ percentile of the EG distribution. If it is lower than all the bootstrapped rates, it is noted to lie at the 0^th^ percentile of the EG distribution.

### Estimation of EG rates in major cancer classification subsets

Each subset is defined as a collection of specific cancer types. Specifically, we use previously established ROSETTA code assignments to group tumor types into adenocarcinomas, malignant melanomas, transitional cell carcinomas, and squamous cell carcinomas for downstream analysis. To compute EG rate and its associated metrics as described above, we subset the incidence vector, total sequenced cases vector, and count matrix to only include counts from the specific cancer types of interest (e.g., adenocarcinoma ROSETTA codes). The subset incidence vector is normalized to one. Then, we estimate EG rates and confidence intervals as described above with these subset matrices.

### Enrichment scores for mutations and assessing their statistical significance

We define a set of genes G that correspond to Tier 1 of the Cancer Gene Census (*30*) (Supplementary File 1). Then, we implement the gene-set enrichment score often used in RNA-Seq analyses (94). Namely, compute phit - the sum of all EG rate estimates for mutations belonging to genes in set G. Then, compute pmiss, defined as the total number of unique mutations in the dataset minus the number of mutations with genes belonging to set G. For each mutation i, compute its score *S*_*i*_ which is the EG rate divided by phit if that mutation comes from a gene belonging to set G. Else, *S*_*i*_ is −1/pmiss. Finally, rank the mutations from highest to lowest EG rate and compute the cumulative sum. Thus, at each rank r, the enrichment score is 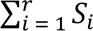. The values of the enrichment score, as in gene-set enrichment analysis, can be plotted against their ranks and visualized.

To assess whether the enrichment score associated with a gene set G, of size n, was statistically significant, we employed a Monte Carlo approach. Namely, we randomly selected n ranks and re-computed the enrichment score. We repeated this process 1000 times resulting in a set of enrichment scores constructed from random gene sets. The p-value was defined as the fraction of resampled enrichment scores greater than the enrichment score constructed from the intended gene set G. A p-value threshold of 0.01 was considered statistically significant.

### Pathways and Gene level analyses

*BRAF* codons were grouped into distinct classes (I, II, III, NC) based on assignment rules from prior work and on domain expertise (JH) (*25, 58–63*). *TP53* codons were grouped into domains (TAD, PRD, DBD, OD, CTD) using codon-domain mappings from the literature (*51*). Further, RAS, PI3K, and WNT pathway genes were curated based on prior work (*4*) (Supplementary Table 1, Supplementary File 1).

### Statistical Analyses

We compute the similarity between two quantities (e.g., NPC and EG rate) using the Pearson correlation coefficient and its associated p-value (*95*). Line-of-best fits are computed by linear regression of the two quantities.

## Data Availability

This paper analyzes existing, publicly available data. Data are accessible on Zenodo at https://zenodo.org/records/10569725. All original code has been deposited at GitHub and is publicly available at https://github.com/aditharun/missense-nonsense-reweighting-mutations. Any additional information required to reanalyze the data reported in this paper is available from the lead contact upon request.

## Acknowledgements

The authors would like to thank Michael Jones, Melinda Tong, Jacob Kim, Eugene Ke, and Meraj Aziz for helpful conversations on the project, methods, and their communication. This work was supported in part by American Cancer Society Research Grant (RSG-25-1434739-01-DMC) to E.C.S., by National Institutes of Health, National Center for Complementary and Integrative Health (DP2AT011327) to E.C.S, and by National institutes of Health, National Cancer Institute (1R01CA276771-01A1) to T.M.

## Contributor Roles

Conceptualization: AA, GM, ES; Data Curation: AA, DL, GM, JH, ES; Formal Analysis: AA, GM, DL; Funding Acquisition: ES; Investigation: AA, DL, GM, ES; Methodology: AA, GM, ES; Project Administration: ES; Resources: ES; Supervision: ES, TM, DH, GW, JH; Visualization: AA, ES; Writing – original draft: AA, ES; Writing – review & editing: AA, TM, JH, ES

---

**Supplementary Figure 1.**
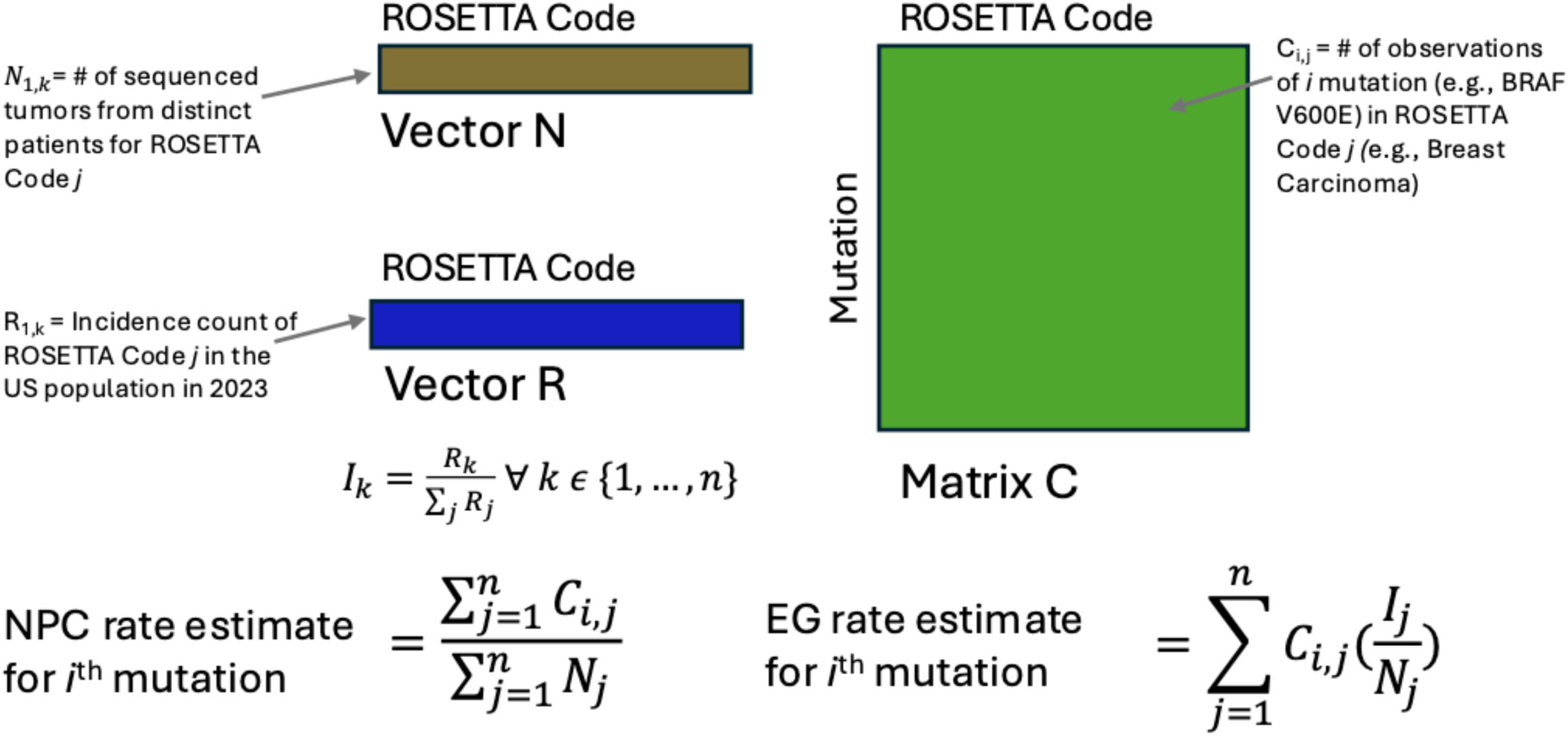
The calculation of epidemio-genomic rates. Schematic describing the reweighting strategy to estimate the epidemio-genomic (EG) rate and naïve pan-cancer (NPC) rate for an individual mutation.

**Supplementary Figure 2.**
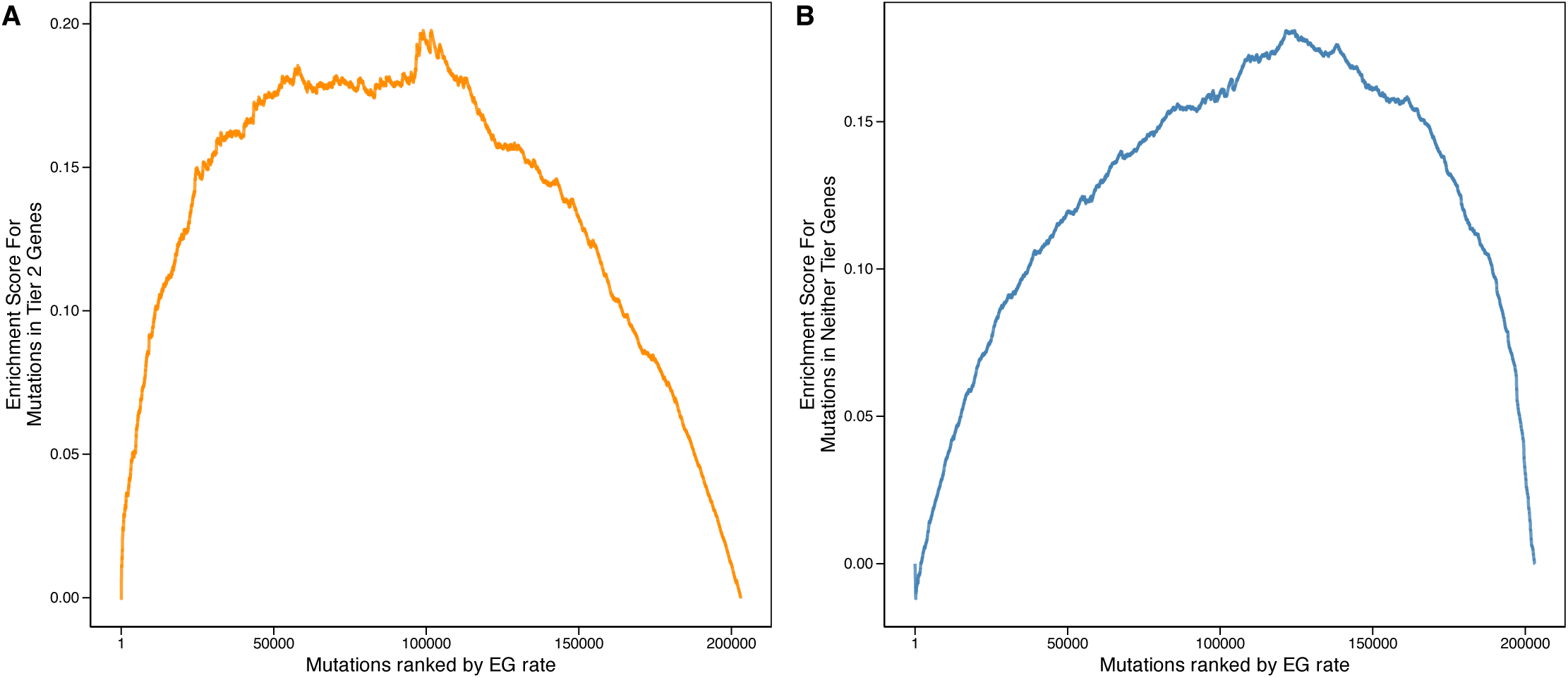
Enrichment of Tier 2 and Neither Tier 1 nor Tier 2 gene sets. A. Enrichment plot for mutations in Tier 2 cancer driver genes among the set of all mutations observed more than once. B. Enrichment plot for mutations in genes that are neither Tier 1 nor Tier 2 cancer driver genes among the set of all mutations observed more than once.

**Supplementary Figure 3.**
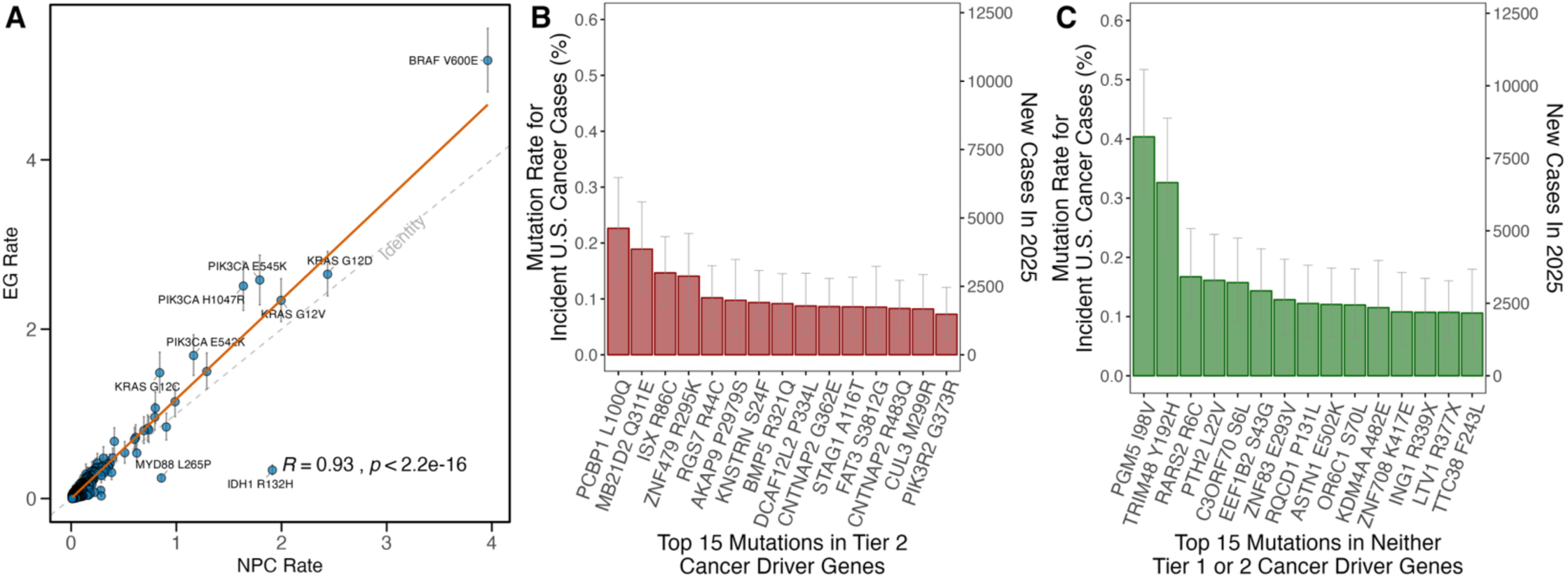
Calculated cancer patient mutation proportions (EG rates) for common missense and nonsense mutations. A. Scatter plot of NPC rate against EG rate with line of best fit shown in red. B. Estimated mutation proportion in incident cases and new patients affected in 2025 for the fifteen most common mutations that occur in genes that are members of Tier 2 of the Cancer Gene Census. C. Estimated mutation proportion in incidence cases (and new patients affected in 2025) for the fifteen most common mutations that occur in genes that are not members of Tier 1 or Tier 2 of the Cancer Gene Census.

**Supplementary Figure 4.**
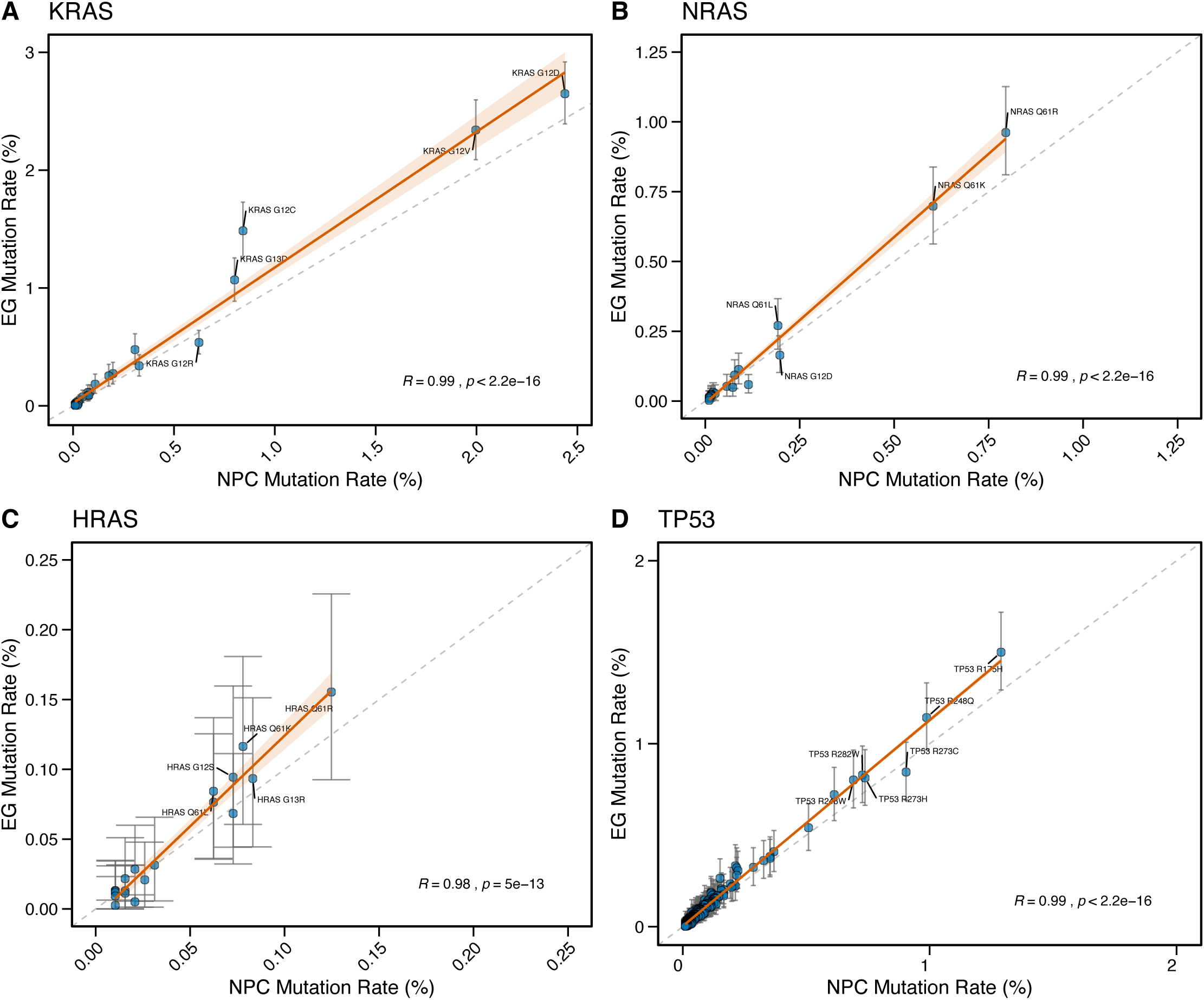
Comparison between EG and NPC rates. Scatter plot for each missense and nonsense mutation observed in (A) *KRAS*, (B) *NRAS*, (C) *HRAS*, and (D) *TP53*. The line of best fit, the 95% confidence interval for the line of best fit, Pearson correlation coefficient and associated p-value are shown. The identity line is shown as grey dashed line.

**Supplementary Figure 5.**
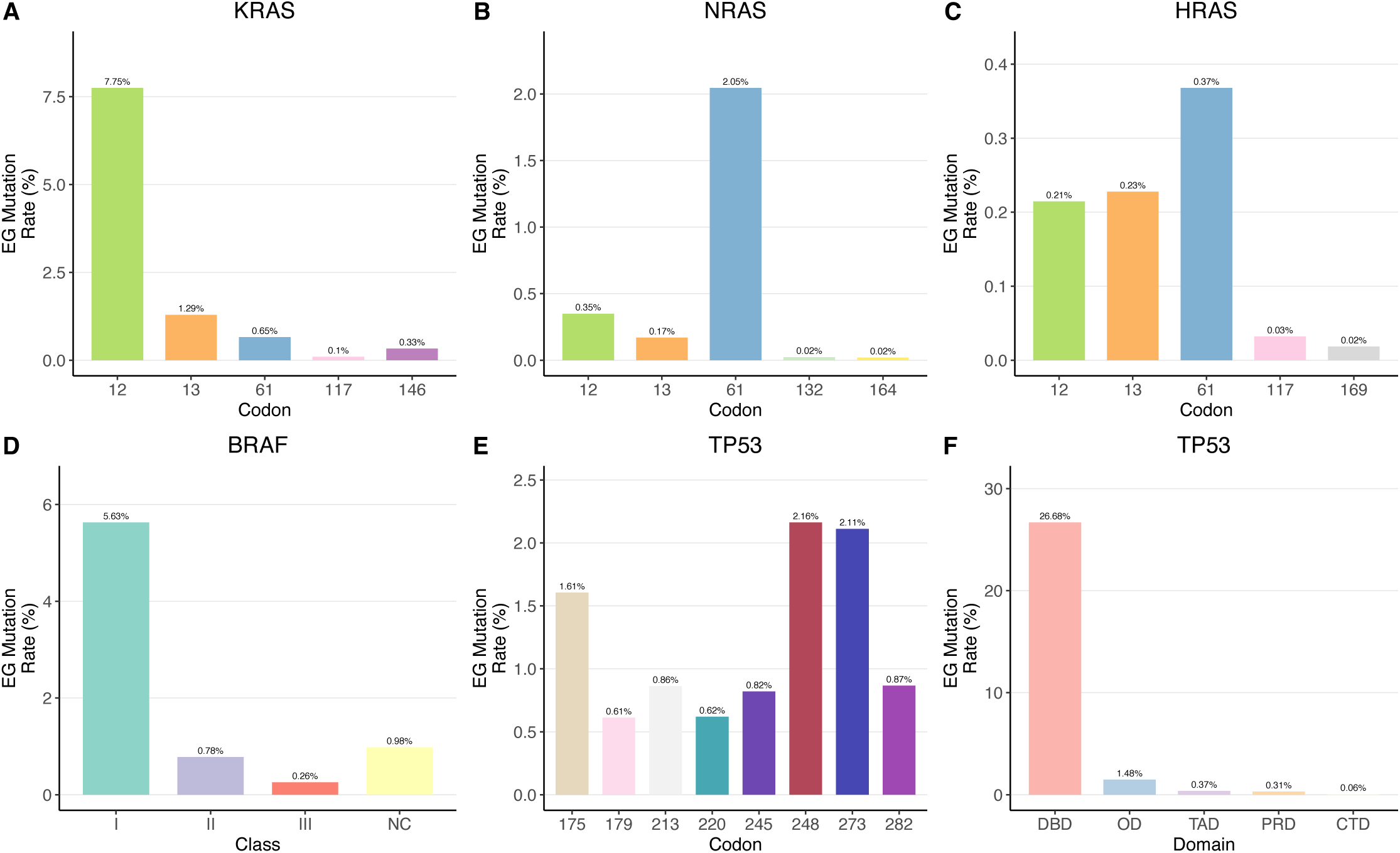
Epidemiogenomic (EG) Mutation Rates of important classes of KRAS, NRAS, HRAS, BRAF, and TP53 mutations. A. Bar chart showing the EG rate for the specified *KRAS* codons in Figure 6A. B. Bar chart showing the EG rate for the specified *NRAS* codons in Figure 6B. C. Bar chart showing the EG rate for the specified *HRAS* codons in Figure 6C. D. Bar chart showing the EG rate for the specified *BRAF* mutation classes in Figure 6D. E. Bar chart showing the EG rate for the specified *TP53* codons in Figure 6E. F. Bar chart showing the EG rate for the specified *TP53* domains in Figure 6F.

**Supplementary Table 1. Catalogue of all recurrent missense and nonsense mutations.** Epidemio-genomic (EG) rate estimates with 95% confidence intervals and naïve pan-cancer (NPC) rate estimates along with EG rate, confidence intervals, and NPC rate for subsets of adenocarcinomas, squamous cell carcinomas, transitional cell carcinomas, and malignant melanomas. Observed number of mutant samples for each tumor type are provided and overall number of US patients affected each year. Estimate of the numerical percentile within the EG bootstrap rate estimates where the NPC rate lies are provided. Binary flags are included for Cancer Gene Census Tier (Tier 1, Tier 2, or neither Tier 1 or Tier 2 consensus cancer driver gene), and pathway (RTK/RAS, PI3K, WNT/β-catenin) status.

**Supplementary File 1. Catalogue of all missense and nonsense mutations.** Epidemio­genomic (EG) rate estimates with 95% confidence intervals and naïve pan-cancer (NPC) rate estimates along with EG rate, confidence intervals, and NPC rate for subsets of adenocarcinomas, squamous cell carcinomas, transitional cell carcinomas, and malignant melanomas. Observed number of mutant samples for each tumor type are provided and overall number of US patients affected each year. Estimate of the numerical percentile within the EG bootstrap rate estimates where the NPC rate lies are provided.

